# The impact of psychological treatment on catastrophization and pharmacological response in chronic migraine: A single-center experience

**DOI:** 10.1101/2024.11.08.24315876

**Authors:** Federica Nicoletta Sepe, Claudia Lanni, Daniele De Michelis, Giacomo Lancia

**Affiliations:** Stroke Unit Department, IRCCS C. Mondino Foundation, Via Mondino, 2, 27100, Pavia, Italy; Headache Clinic, Department of Neurology, Azienda Sanitaria SS. Antonio e Biagio e Cesare Arrigo, Via Venezia, 16, 15121, Alessandria, Italy; Department of Basic and Applied Sciences for Engineering (SBAI), University of Rome “La Sapienza”, Via Antonio Scarpa, 4. 00161, Roma

**Keywords:** Psychological treatment, Chronic migraine, Catastrophization, Pain Catastrophizing Scale

## Abstract

**Background:** We aimed to assess the impact of pain catastrophizing, measured using the Italian version of the Pain Catastrophizing Scale (PCS), on the clinical response of patients with chronic migraine to anti-CGRP monoclonal antibodies combined with a multidisciplinary approach, including psychological treatment.

**Methods:** 25 Outpatients from SS. Antonio e Biagio e Cesare Arrigo headache clinic randomly assigned to receive Galcanezumab, Erenumab, or Fremanezumab. Their clinical response was evaluated over six months using various measures, including reducing the number of days with migraine per month, and quality of life using Headache Impact Test (HIT 6), MIgraine Disability Assessment Score questionnaire (MIDAS), and Beck’s Inventory Scale (BDI II) scales to assess comorbid depression.

**Results:** We established a strong correlation between HIT 6 and PCS, with coefficients of 0.81 and 0.88 at *T*_1_ and *T*_2_, respectively. Furthermore, we found no significant correlation between PCS and the other scales, such as MIDAS, as with any pharmacological therapies.

**Conclusion:** This study aims to clearly define the impact of a multidisciplinary approach including a psychological follow-up on a particular clinical phenotype of chronic migraines and their tendency to catastrophize, but more extended data are needed.

## 1 Introduction

The PCS is a 13-item self-report measure of catastrophizing in pain and a well-validated measure of maladaptive thinking patterns related to pain (Monticone et al., 2012). The PCS is composed of 3 subscales: rumination (e.g., “I can’t stop thinking about how much it hurts”), magnification (e.g., “I worry that something serious may happen”), and helplessness (e.g., “There is nothing I can do to reduce the intensity of the pain”) (Monticone et al., 2012). People affected by chronic headaches, defined according to the 3rd edition of the International Classification of Headache Disorders (ICHD III) (Arnold, 2018), represent a consistent group of patients suffering from chronic pain. Giving the best medical response to those patients, affording the considerable economic costs and psychological burden related to chronic pain, is a huge healthcare challenge. Since the 90s, researchers focused on the role of anxiety and mood disorders as comorbidities in migraine, without describing the specific role of a patient’s personality in the processing of pain (Pelzer et al., 2023a; Lipton et al., 2023; Klonowski et al., 2022). Catastrophizing can be considered a maladaptive cognitive response to a painful stimulation that influences negatively pain perception (Mathur et al., 2016). In high-frequency migraine patients, pain-related activity in the white matter structure of the insula correlates with pain catastrophizing and migraine severity (Kocakaya et al., 2023). Furthermore, in patients with medical overuse (MOH), a higher total PCS score was associated with decreased grey matter density in precentral and inferior temporal gyrus as an increased resting-state functional connectivity between middle temporal gyrus and cerebellum (Christidi et al., 2020). An abnormal reward mechanism dopamine-mediated can explain these structural changes: high-frequency migraine attacks induced sustained increases in dopaminergic trafficking that override homeostatic feedback control. This massive dopaminergic tone in reward could lead motivation and learning centres to actuate abnormal coping strategies such as catastrophizing for pain (Christidi et al., 2020; Maizels et al., 2012). Even if the role of pain-related cognitive processes and emotional state on pain-related disability is well established (Senturk et al., 2023a), how catastrophizing can influence therapeutical response in the calcitonin-gene related peptide (CGRP-mAbs) antibodies era is still debated. Interestingly, refractory migraine patients to CGRP-mAbs showed higher baseline PCS scores (Alvarez-Astorga et al., 2021) representing an independent negative predictor to CGRP-mAbs response (Silvestro et al., 2021; Mitsikostas et al., 2014).

This study aims to assess the potential impact of pain catastrophizing on the clinical response to CGRP-mAbs in a real-life setting at a tertiary headache centre in Northern Italy.

## 2 Materials and Methods

### 2.1 Ethics Committee

The local Clinical Research Ethics Committee approved the study (number of approval ASO.NEURO.21.02), and all patients signed informed consent forms

### 2.2 Study population

In this monocentric observational study, we enrolled 25 consecutive outpatients who visited the “SS. Antonio e Biagio e Cesare Arrigo” headache clinic between July 2021 and 2023. These were diagnosed with chronic migraine, with or without medication overuse, based on the ICDH III criteria. They were randomly assigned to receive one of three medications — Galcanezumab (120mg), Erenumab (140mg), or Fremanezumab (225mg) — following the local and EAN guidelines. The therapeutical response was assessed using two criteria: a > 50% reduction in the frequency of number of days with migraine per month and a decrease in disability using the MIDAS (Stewart et al., 2001a) and the HIT 6 (Rendas-Baum et al., 2014) according to local and EAN guidelines. We also evaluated comorbid depression using the BDI II(BDI II > 13) (Schotte et al., 1997a). Patients were assessed at the beginning of therapy (*T*_0_) and again at three months (*T*_1_) and six months (*T*_2_) after the start of treatment. Specialists in neurology and psychology treated each patient, and those with medical, psychiatric, or cognitive limitations, as well as those with chronic pain conditions or a history of substance abuse, were excluded from the study. The sociodemographic and clinical characteristics of patients are reported in Table 1.

**Table 1:** Demographic and clinical background details encompassing the entire patient cohort. Blank cells represent missing values. An X indicates the assignment of antibodies.

| Sex | Age | Ere<br>nu<br>mab | Fre<br>ma<br>ne<br>zu<br>mab | Gal<br>ca<br>ne<br>zu<br>mab | MID<br>AS<br>($T_0$) | HIT<br>6<br>($T_0$) | BDI<br>II<br>($T_0$) | PCS<br>($T_0$) | Help<br>ness<br>($T_0$) | Mag<br>ni<br>fi<br>ca<br>tion<br>($T_0$) | Ru<br>mi<br>na<br>tion<br>($T_0$) | MID<br>AS<br>($T_1$) | HIT<br>6<br>($T_1$) | BDI<br>II<br>($T_1$) | PCS<br>($T_1$) | Help<br>ness<br>($T_1$) | Mag<br>ni<br>fi<br>ca<br>tion<br>($T_1$) | Ru<br>mi<br>na<br>tion<br>($T_1$) | MID<br>AS<br>($T_2$) | HIT<br>6<br>($T_2$) | BDI<br>II<br>($T_2$) | PCS<br>($T_2$) | Help<br>ness<br>($T_2$) | Mag<br>ni<br>fi<br>ca<br>tion<br>($T_2$) | Ru<br>mi<br>na<br>tion<br>($T_2$) |
| --- | --- | --- | --- | --- | --- | --- | --- | --- | --- | --- | --- | --- | --- | --- | --- | --- | --- | --- | --- | --- | --- | --- | --- | --- | --- |
| F | 50-54 |  | X |  | 87 | 72 | 18 | 28 | 7 | 7 | 14 | 44.0 | 67.0 | 12.0 | 27.0 | 11.0 | 2.0 | 14.0 | 30.0 | 73.0 | 11.0 | 24.0 | 6.0 | 4.0 | 14.0 |
| F | 25-29 |  | X |  | 75 | 72 | 23 | 42 | 22 | 2 | 18 | 40.0 | 44.0 | 30.0 | 7.0 | 1.0 | 4.0 | 2.0 | 18.0 | 62.0 | 9.0 | 31.0 | 15.0 | 0.0 | 16.0 |
| F | 70-74 |  | X |  | 90 | 76 | 27 | 39 | 17 | 2 | 20 | 15.0 | 67.0 | 21.0 | 35.0 | 16.0 | 2.0 | 17.0 | 34.0 | 57.0 | 15.0 | 17.0 | 3.0 | 2.0 | 12.0 |
| F | 55-59 |  | X |  | 63 | 71 | 12 | 42 | 18 | 4 | 20 |  |  |  |  |  |  |  |  |  |  |  |  |  |  |
| F | 20-24 |  | X |  | 160 | 65 | 30 | 33 | 15 | 2 | 16 | 43.0 | 60.0 | 7.0 | 29.0 | 11.0 | 0.0 | 18.0 | 58.0 | 59.0 | 4.0 | 22.0 | 9.0 | 0.0 | 13.0 |
| F | 45-49 |  | X |  | 76 | 44 | 10 | 26 | 8 | 0 | 18 | 80.0 | 54.0 | 8.0 | 24.0 | 8.0 | 0.0 | 16.0 |  |  |  |  |  |  |  |
| F | 55-59 | X |  |  | 68 | 60 | 14 | 23 | 11 | 2 | 10 | 60.0 | 60.0 | 6.0 | 20.0 | 6.0 | 1.0 | 13.0 | 30.0 | 62.0 | 13.0 | 18.0 | 6.0 | 2.0 | 10.0 |
| F | 50-54 | X |  |  | 57 | 63 | 11 | 22 | 7 | 2 | 13 | 13.0 | 40.0 | 1.0 | 12.0 | 1.0 | 0.0 | 11.0 | 16.0 | 57.0 | 6.0 | 15.0 | 3.0 | 0.0 | 12.0 |
| F | 45-49 | X |  |  | 119 | 68 | 39 | 26 | 12 | 0 | 14 | 70.0 | 68.0 | 38.0 | 25.0 | 14.0 | 0.0 | 11.0 | 29.0 | 56.0 | 40.0 | 13.0 | 3.0 | 1.0 | 9.0 |
| F | 55-59 | X |  |  | 69 | 72 | 12 | 43 | 21 | 2 | 20 |  |  |  |  |  |  |  |  |  |  |  |  |  |  |
| F | 40-44 | X |  |  | 97 | 67 | 17 | 22 | 8 | 0 | 14 | 29.0 | 50.0 | 15.0 | 16.0 | 4.0 | 2.0 | 10.0 | 9.0 | 36.0 | 6.0 | 3.0 | 0.0 | 1.0 | 2.0 |
| M | 65-69 | X |  |  | 30 | 63 | 23 | 32 | 12 | 0 | 20 | 30.0 | 46.0 | 11.0 | 11.0 | 1.0 | 1.0 | 9.0 | 12.0 | 36.0 | 11.0 | 8.0 | 0.0 | 1.0 | 7.0 |
| F | 65-69 | X |  |  | 12 | 60 | 3 | 24 | 14 | 2 | 8 | 11.0 | 46.0 | 9.0 | 15.0 | 6.0 | 2.0 | 7.0 |  |  |  |  |  |  |  |
| F | 55-59 |  |  | X | 56 | 67 | 22 | 39 | 17 | 5 | 17 | 0.0 | 36.0 | 6.0 | 10.0 | 1.0 | 0.0 | 9.0 | 0.0 | 40.0 | 13.0 | 10.0 | 3.0 | 2.0 | 5.0 |
| M | 50-54 |  |  | X | 56 | 64 | 9 | 26 | 9 | 6 | 11 | 10.0 | 52.0 | 3.0 | 8.0 | 2.0 | 1.0 | 5.0 | 4.0 | 40.0 | 1.0 | 10.0 | 2.0 | 1.0 | 7.0 |
| F | 35-39 |  |  | X | 26 | 62 | 14 | 30 | 12 | 2 | 14 | 20.0 | 44.0 | 11.0 | 10.0 | 3.0 | 1.0 | 9.0 | 22.0 | 40.0 | 12.0 | 5.0 | 1.0 | 2.0 | 8.0 |
| F | 55-59 |  |  | X | 20 | 65 | 13 | 27 | 11 | 4 | 12 | 4.0 | 32.0 | 5.0 | 9.0 | 1.0 | 1.0 | 5.0 | 2.0 | 34.0 | 1.0 | 11.0 | 2.0 | 1.0 | 7.0 |
| M | 45-49 |  |  | X | 36 | 69 | 7 | 38 | 18 | 4 | 16 |  |  |  |  |  |  |  |  |  |  |  |  |  |  |
| F | 60-64 |  |  | X | 87 | 62 | 11 | 35 | 15 | 4 | 16 | 12.0 | 62.0 | 10.0 | 26.0 | 10.0 | 3.0 | 13.0 |  |  |  |  |  |  |  |
| F | 50-54 |  |  | X | 96 | 76 | 12 | 30 | 12 | 2 | 16 |  |  |  |  |  |  |  |  |  |  |  |  |  |  |

**Table 2:** Descriptive statistics with Standardized moments for PCS, HIT 6, MIDAS, and BDI II at times *T*_0_, *T*_1_, and *T*_2_.

|  | Mean | Variance | Skewness | Kurtosis | Tailing |
| --- | --- | --- | --- | --- | --- |
| PCS $T_0$ | 30 | 97 | -0.58 | 3.75 | -6.66 |
| PCS $T_1$ | 20 | 49 | 0.11 | 1.95 | 0.38 |
| PCS $T_2$ | 15 | 44 | 0.19 | 2.94 | 1.81 |
| HIT6 $T_0$ | 65 | 21 | 0.45 | 2.55 | 2.96 |
| HIT6 $T_1$ | 55 | 93 | -0.43 | 2.26 | -2.33 |
| HIT6 $T_2$ | 54 | 110 | -0.23 | 2.89 | -0.63 |
| MIDAS $T_0$ | 67 | 1135 | 0.79 | 3.43 | 6.29 |
| MIDAS $T_1$ | 29 | 349 | 0.23 | 2.81 | 2.38 |
| MIDAS $T_2$ | 26 | 557 | 1.32 | 3.42 | 9.11 |
| BDI II $T_0$ | 20 | 92 | 0.38 | 2.62 | 2.34 |
| BDI II $T_1$ | 14 | 118 | 1.05 | 3.16 | 5.81 |
| BDI II $T_2$ | 11 | 75 | 1.38 | 7.48 | 23.91 |

## 3 Statistical Analysis

As a primary outcome, we investigated the correlation between PCS and HIT 6 at *T*_0_, *T*_1_, and *T*_2_ using Spearman’s correlation test; see appendix A. We also examined the association between PCS and MIDAS and PCS and BDI II at these time points. A significance level of 5% was set for all hypotheses; with a Bonferroni correction, we set the p-value threshold at 0.00625.

We also assessed the reduction in MIDAS and HIT 6 using Wilcoxon’s test, with the significance level adjusted to 0.00625; see appendix B.

Using the Jaccard Index, we investigated the similarity of subgroups of patients showing severe catastrophization (PCS > 30) as headache-related disability, considering HIT 6 > 50and MIDAS > 11 separately. Depression at *T*_0_ was controlled using BDI II (> 13) as the antibody assigned; see C.

Using a logit approach, we explored the relationship between PCS at *T*_2_ and baseline information at *T*_0_. The outcome variable was defined as a relative reduction of MIDAS > 50% between *T*_0_ and *T*_2_. Permutation Importance identified the most impactful features for improving quality of life; see appendix D.

We focused on the primary outcome of PCS and HIT 6 correlation through Spearman’s test to determine the sample size. Therefore, we utilized the method proposed by May and Looney (2020), setting a significance level of 0.0125 (adjusted for Bonferroni correction) and a power of 0.8; see appendix E. We estimated a sample size of 21, adjusted to 25 for a 15% dropout rate.

We used a simple randomization scheme to assign antibody therapy, ensuring equal assignment probabilities. Despite a higher number of patients receiving Galcanezumab, the chi-squared test confirmed no significant distribution difference among patients at a 5% significance level; see appendix F.

## 4 Results

The null hypothesis of no correlation between PCS and HIT 6 was rejected with p-values of 0.001 at *T*_0_, *T*_1_, and *T*_2_. Spearman’s coefficients were 0.65, 0.81, and 0.88 at *T*_0_, *T*_1_, and *T*_2_, respectively. These results describe a strong and time-dependent correlation between PCS e HIT 6 scores. For PCS and MIDAS, the null hypothesis could not be rejected at *T*_0_ and *T*_1_ (p-value: 0.35 and 0.07) but was rejected at *T*_2_ (p-value < 0.001), with correlation coefficients of 0.81 and 0.68. So, as expected, a reduction in the tendency to catastrophize will directly impact on quality of life and number of attacks per month measured by MIDAS scale.

Significant differences were found for all scale scores between *T*_0_ and *T*_1_: PCS (p-value < 0.001), HIT 6 (p-value < 0.001), and MIDAS (p-value <0.001). Significant differences were also observed between *T*_0_ and *T*_2_: PCS (p-value <0.001), HIT 6 (p-value <0.001), and MIDAS (p-value <0.001). Figure 1 provides a detailed breakdown.

**Figure 1.**
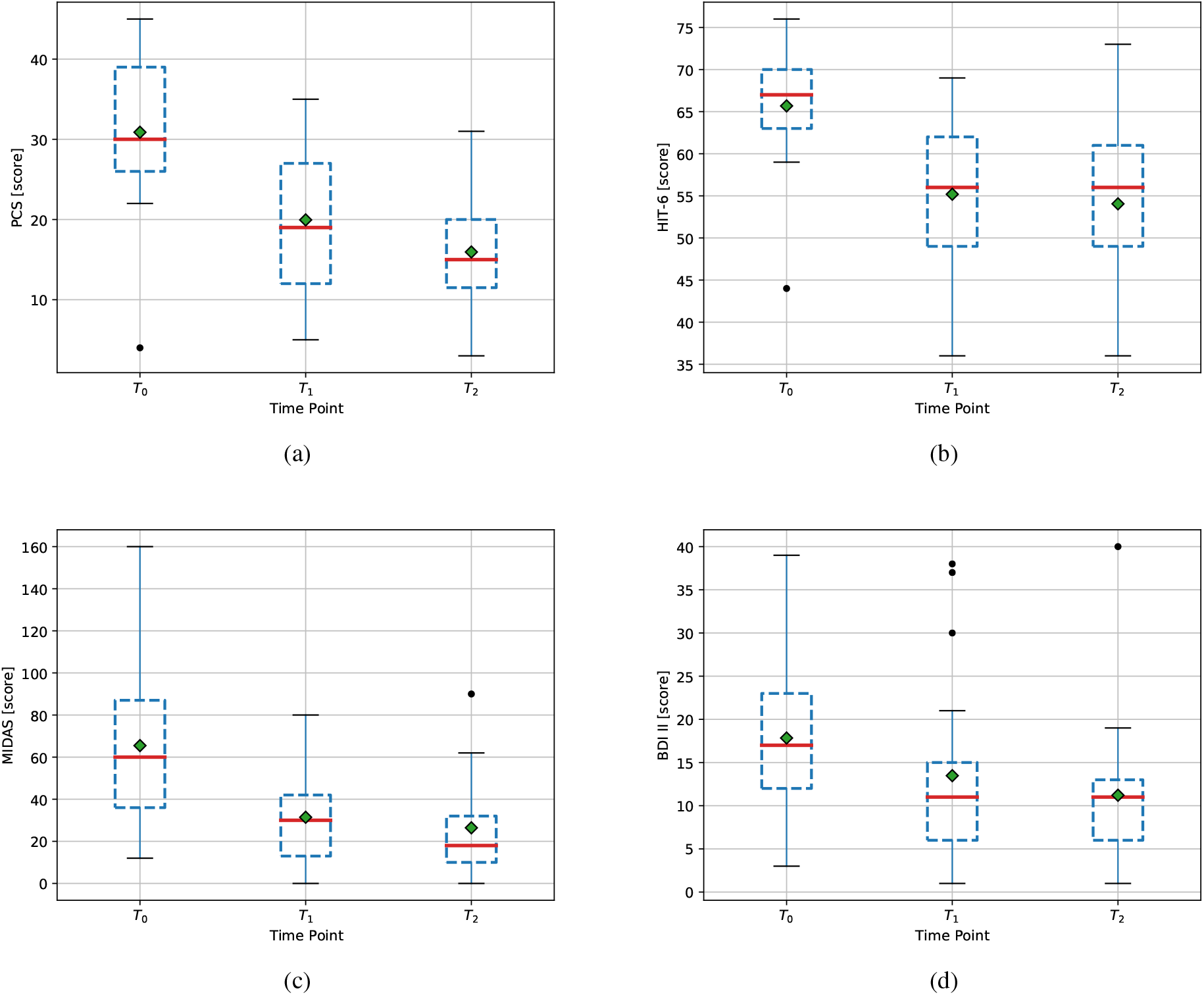
Panel with box plots for (a) PCS, (b) HIT6, (c) MIDAS, and (d) BDI II scales. The interquartile range is enclosed within the box, while the red dashed line is the median value. The green diamond is the average value. Black dots denote outliers.

At the beginning of the study (*T*_0_), the Jaccard Index indicated a 47% agreement between the reduction of severe catastrophization scores and the other scales. This result indicates that 47% of patients with a high PCS score responded positively (the highest reduction in monthly migraine days) to both psychological and pharmacological treatments. This reduction occurred regardless of the type of antibody, except for a slight correlation with Galcanezumab treatment. Our model suggests that comorbid depression and antidepressant therapy at *T*_0_ have no influence.

The logit-based model had an AUROC of 0.75 (95% CI 0.73-0.78), i.e., one patient has a probability of 75% to have a good clinical response at six months from the beginning of the therapy. The logit-based model was inspected through the permutation importance method, revealing that age, Galcanezumab, and PCS score at *T*_0_ are the main factors in the clinical response, with importance scores of 0.29, 0.17, and 0.28 (95% CI 0.26-0.32; 0.15-0.18; 0.26-0.30), respectively.

Missing values resulted from right censoring, affecting six patients at either *T*_1_ or *T*_2_. We handled the missing data by analyzing all available data; we excluded patients whose therapy was interrupted and for whom no further information was available.

## 5 Discussion

Our hypothesis posits that rumination, as a self-psychological condition, may directly influence the clinical efficacy of CGRP antibodies, particularly in cases deemed refractory. We propose that a multidisciplinary approach incorporating psychological support can significantly improve clinical response, especially for those exhibiting an inadequate response to treatment. However, identifying the specific patient phenotype that will benefit the most from this approach remains challenging. Surprisingly, patients with the best clinical response appear to be female and younger, with higher PCS scores at baseline, regardless of comorbid depression. Galcanezumab seems to be more effective. There is a significant time-dependent correlation between the reduction in PCS and HIT 6 scores, particularly when longer medical and psychological therapies have been administered. This is independent of the number of attacks reported as of antidepressant previous therapies or depression. This suggests that a holistic treatment approach - combining pharmacological and psychological interventions - is more likely to improve the quality of life for chronic migraine patients. In a recent study, the HIT 6 score was found to have a weak correlation with PCS scores (Kim et al., 2021). However, no investigation was made into the correlation between any therapy and these variables. Another multicentric study also found that helplessness and anxiety are linked to the social quality of life of migraine patients when compared to controls. However, no correlations to ABS treatment were identified (Senturk et al., 2023b). We agree with previous theses that pain catastrophizing may indicate a clinical phenotype with heightened expression of altered central sensitization and consequential coping mechanisms leading to an overall decrease in quality of life (Sciruicchio et al., 2019). Identifying the various factors that drive the progression of chronic migraine is crucial to developing effective prevention strategies (Driessen et al., 2022). Other real-world studies demonstrated that CGRP antibodies were able to exert an excellent clinical response across subgroups of migraine patients with comorbid psychological traits, mainly anxiety, and depression (Smitherman et al., 2020; Pelzer et al., 2023b). Unfortunately, this study has several limitations, mainly due to its observational monocentric design: the small sample size, the absence of a control group, and the short observation time. Further data of a more extensive nature is imperative to substantiate our findings.

## 6 Conclusion

Treatment-related and individual factors contribute to the development of chronic migraine. A comprehensive approach is essential for individuals, encompassing both pharmacological and non-pharmacological interventions, as well as the management of behavioural and psychological factors. The development of personalized tools for predicting chronification represents a significant research priority. This study aims to delineate a clinical phenotype of chronic migraine patients characterized by a propensity to catastrophize. It advocates for a combined therapeutic strategy utilizing CGRP monoclonal antibodies alongside psychological counselling to enhance quality of life. Further investigation is necessary to assess the clinical implications of this integrative approach.

### A Spearman’s test

We utilized Spearman’s test (Spearman, 1961) to investigate the correlation between catastrophization (measured with PCS) and MIDAS, HIT 6, and BDI II scales. Specifically, we studied correlations at time *T*_0_ (starting of antibody treatment), *T*_1_ (3 months later), and *T*_2_ (six months later). For each couple of variables (i.e., PCS vs. MIDAS, PCS vs. HIT 6, and PCS vs. BDI II)), we conducted a separate analysis.

Moreover, through the same approach, we studied the correlation level between the PCS subscales such as Helpness, Magnification, and Rumination, and the other scales mentioned above (i.e., MIDAS, HIT 6, and BDI II). Also, the correlation levels between the scales utilized and the antidepressant treatment at *T*_0_ were considered.

The significance level *α* was set to 5%. However, the comparison of tests accomplished at the same time required the significance level to be adjusted in order to avoid the risk of a Type I Error. Thus, we utilized the Bonferroni correction (Dunn, 1961), i.e., 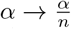, with *n* the number of tests involved. Note that we are interested in testing correlation for two couples of variables along three distinct time points; so we have *n* = 8. As a result, the adjusted significance level is 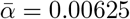.

For completeness, we report in Figures 2-4 the complete matrixes of the Spearman’s coefficients for each variable in our posses along with the p-values matrix; each matrix shows correlation levels at one of the three time-points *T*_0_, *T*_1_, and *T*_2_. The descriptive statistics of the scale scores are shown in Figure 2.

**Figure 2.**
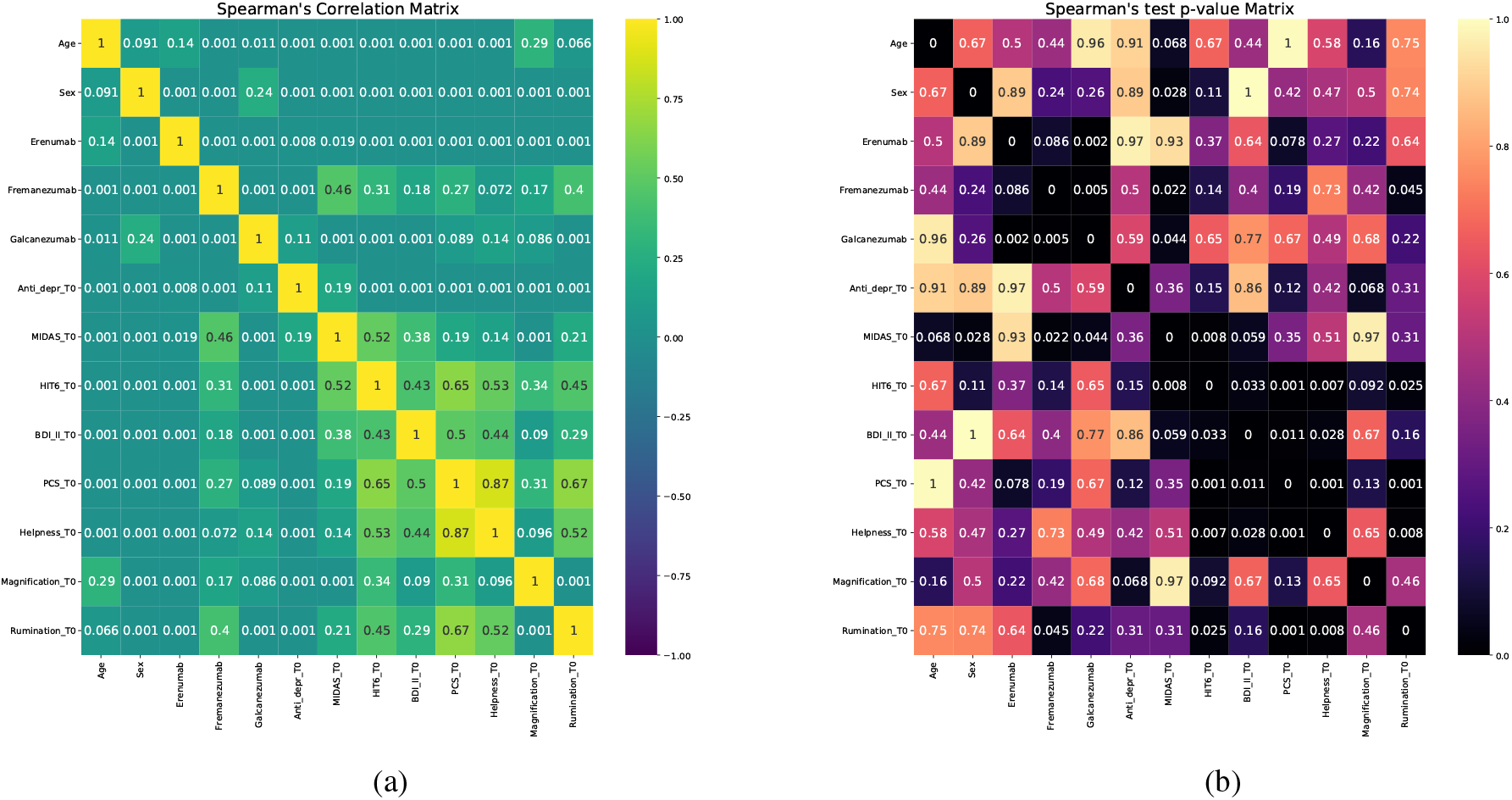
Sperman’s correlation test at T0. (a) Spearman’s correlation coefficients and (b) p-values.

**Figure 3.**
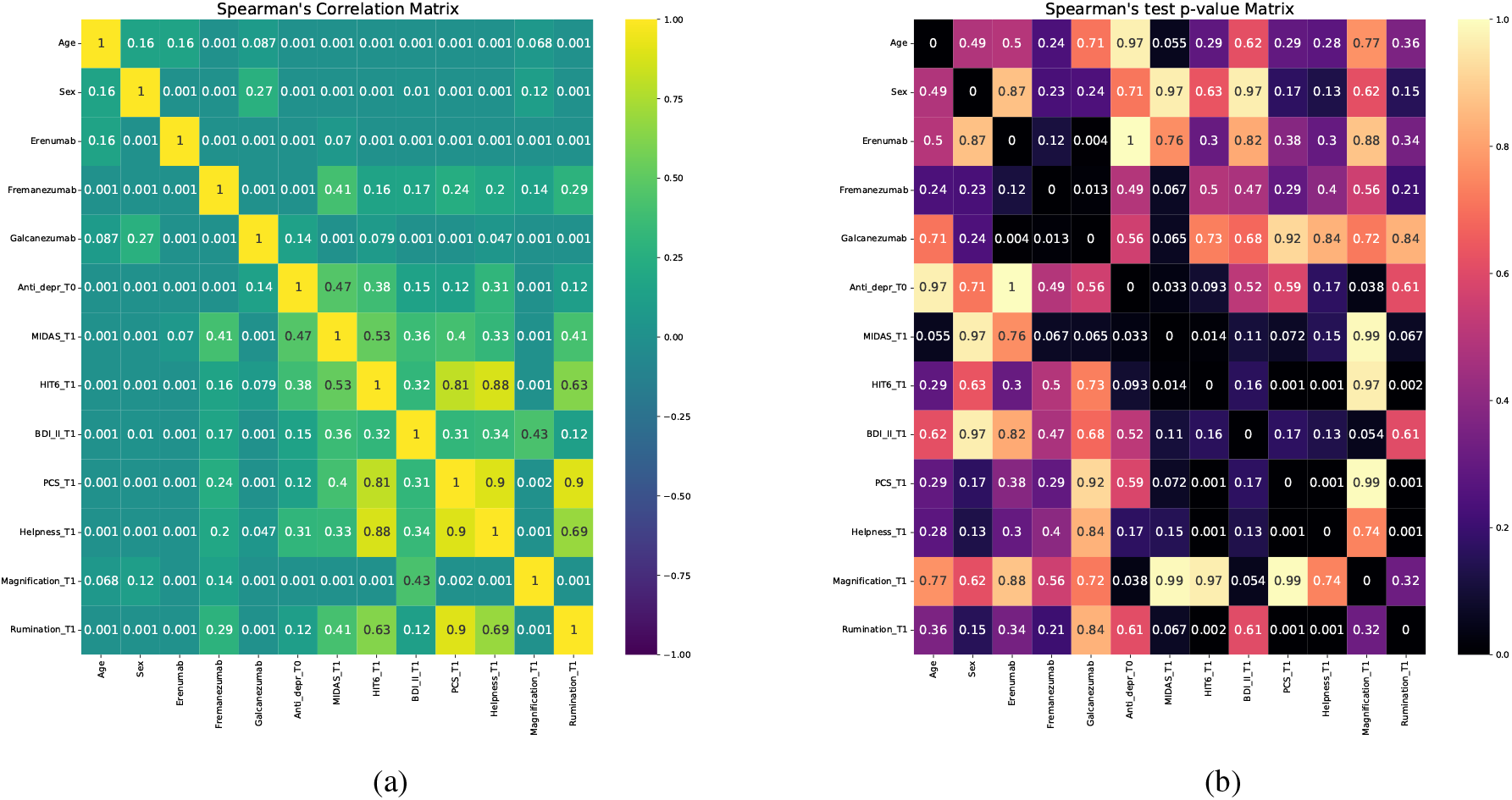
Sperman’s correlation test at *T*_1_. (a) Spearman’s correlation coefficients and (b) p-values.

**Figure 4.**
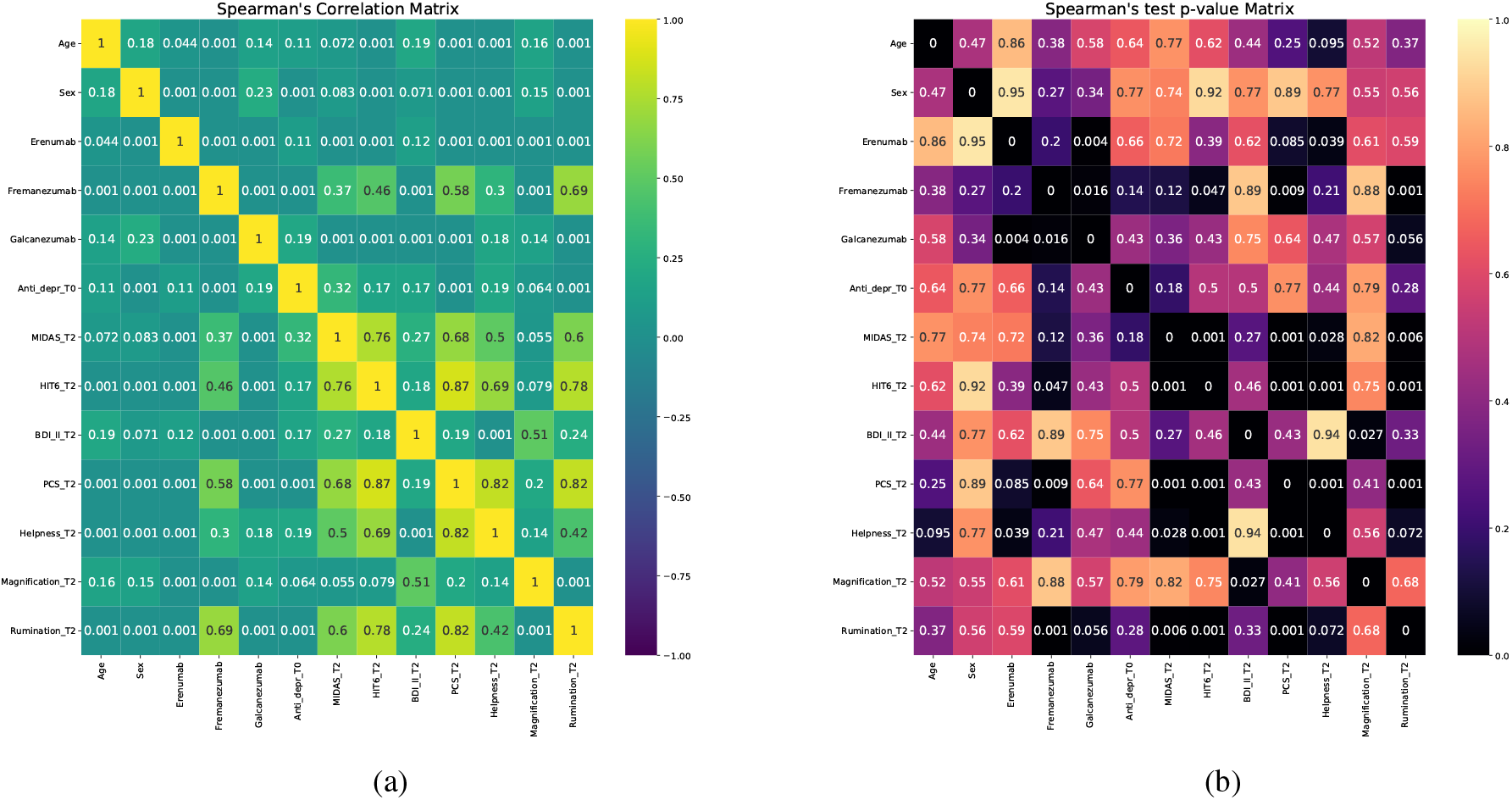
Sperman’s correlation test at *T*_2_. (a) Spearman’s correlation coefficients and (b) p-values.

### B Wilcoxon’s Test

We employed Wilcoxon’s signed-rank test (Conover, 1999; McDonald, 2014) to assess significant differences in scale scores between *T*_0_ and subsequent time points, namely *T*_1_ and *T*_2_. The scale considered is the same utilized in appendix A. Specifically, we conducted one-tailed tests for each pain scale to test the null hypothesis of no change in scale scores at *T*_1_ and *T*_2_ compared to *T*_0_. The alternative hypothesis selected was a decrease in scale scores at *T*_1_ and *T*_2_ compared to *T*_0_.

### C Jaccard Index

The Jaccard Index Chung et al. (2019) was employed to quantify the proportion of patients who exhibited both severe catastrophization and severe migraine-induced disability compared to those who did not.

To assess this relationship we defined a subgroup of patients characterized by severe catastrophization (PCS > 30) and severe headache-related disability (HIT 6 > 50, MIDAS > 11). Thus, our primary goal was to determine whether patients prone to catastrophizing also experience severe disability due to migraines, and vice versa. We assessed the Jaccard Index across both the entire population and specific subpopulations derived through a stratification process. Stratification was performed based on either the antibody treatment received at *T*_0_ or by verifying the presence of comorbid depression (BDI II > 13, minimal depression).

To provide a clearer background about the Jaccard Index in this context, consider the following example. Let *n* denote the generic patient. Define 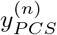 as a dichotomous variable that takes the value 1 if the patient’s PCS score is greater than 30, and 0 otherwise. Similarly, let 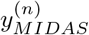 be a dichotomous variable that takes the value 1 if the patient’s MIDAS score is greater than 11, and 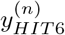 take the value 1 if the patient’s HIT 6 score is greater than 50. Also, let’s consider the sets

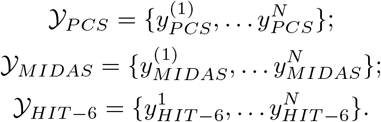

We denoted by superscript one patient within a generic set containing *N* individuals.

The Jaccard index for two sets *A* and *B* of binary items is denied as

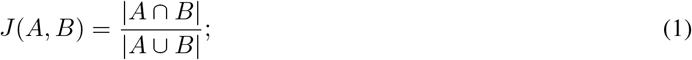

The Jaccard index measures similarity between binary sample sets, defined as the number of items with equal binary labels over the number of items considered.

Therefore, *J*(𝒴_*PCS*_, 𝒴_*MIDAS*_) informs about the percentage of patients which show either PCS > 30 and MIDAS > 11 or PCS < 30 and MIDAS < 11. For both *T*_0_ and *T*_2_, we evaluated *J*(𝒴_*PCS*_, *𝒴*_*MIDAS*_), *J*(𝒴_*PCS*_, *𝒴*_*HIT* −6_), and *J*(𝒴_*HIT* −6_, 𝒴_*MIDAS*_). Results are shown in Tables 3 and 4.

**Table 3:**
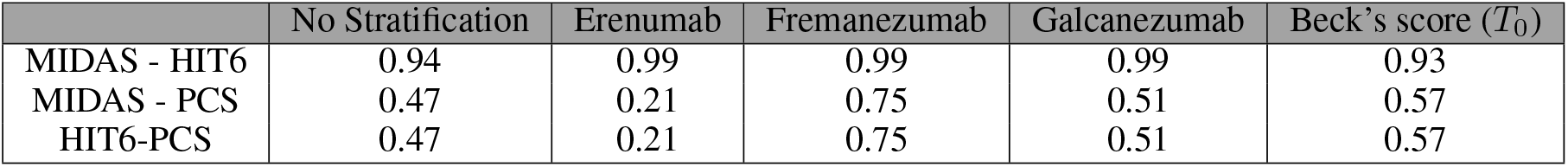
Average Jaccard scores at time *T*_0_ for PCS, HIT6, and MIDAS. Each column reports average association indexes for each stratification (i.e., antibody or binarized Beck’s score at *T*_0_)

**Table 4:** Average Jaccard scores at time *T*_2_ for PCS, HIT6, and MIDAS. Each column reports average association indexes for each stratification (i.e., antibody or binarized Beck’s score at *T*_0_)

| | No Stratification | Erenumab | Fremanezumab | Galcanezumab | Beck's score ($T_0$) |
| --- | --- | --- | --- | --- | --- |
| MIDAS - HIT6 | 0.52 | 0.61 | 0.99 | 0.03 | 0.56 |
| MIDAS - PCS | 0.05 | 0.01 | 0.25 | 0.01 | 0.05 |
| HIT6-PCS | 0.47 | 0.01 | 0.25 | 0.51 | 0.05 |

### D Logit-based approach

Using a logit approach involves modelling the relationship between patients’ quality of life at *T*_2_ and their baseline information at *T*_0_. The logistic regression model is typically used when the outcome variable is binary or categorical. In this case, the outcome variable *Y* is defined based on the relative reduction of MIDAS by more than 50% between *T*_0_ and *T*_2_. This can be represented as:

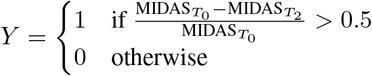

Here, 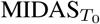 and 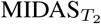 denote the MIDAS scores at baseline *T*_0_ and follow-up *T*_2_, respectively.

Permutation Importance (Breiman, 2001) is a technique used to identify the most impactful features in a predictive model. It works by evaluating the change in model performance (e.g., accuracy, AUC) when the values of a feature are randomly permuted while keeping other features unchanged. The decrease in model performance after permutation indicates the importance of that feature in predicting the outcome.

The logit model is a type of regression analysis used to predict the probability of a binary outcome based on one or more predictor variables. In practice, the probability that patients reduce significantly of 50% the value of MIDAS is given by

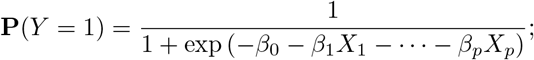

where *β*_0_ is the intercept, *β*_1_, …, *β*_*p*_ are the coefficients for predictors *X*_1_ … *X*_*p*_.

We used a 250-repeated 4-fold stratified cross-validator to evaluate the Area Under the Curve (AUC) and Brier’s Score (BS) to assess the model’s effectiveness. When utilizing the Permutation Importance method, we determined the resilience of each covariate by assigning importance scores based on the average difference of AUC just before and after permutating a covariate.

Note that with the term 250-repeated 4-fold stratified cross-validation approach we refer to the process we utilized to validate the model. It consists of repeating 250 times a 4-fold split into training and test datasets; usually, 3 folds are utilized to train the model, while the leftover is involved in the validation process. The model is therefore trained and tested through 250 different random configurations derived from the sample population. This method ensured robust assessment and minimized overfitting by repeatedly splitting the data into training and testing sets.

To determine the importance of each covariate, we used the Permutation Importance method. This involved calculating the importance scores by measuring the average change in AUC before and after permuting each covariate; see Figure 5 This approach allowed us to assess the resilience and significance of each covariate in predicting the outcome.

**Figure 5.**
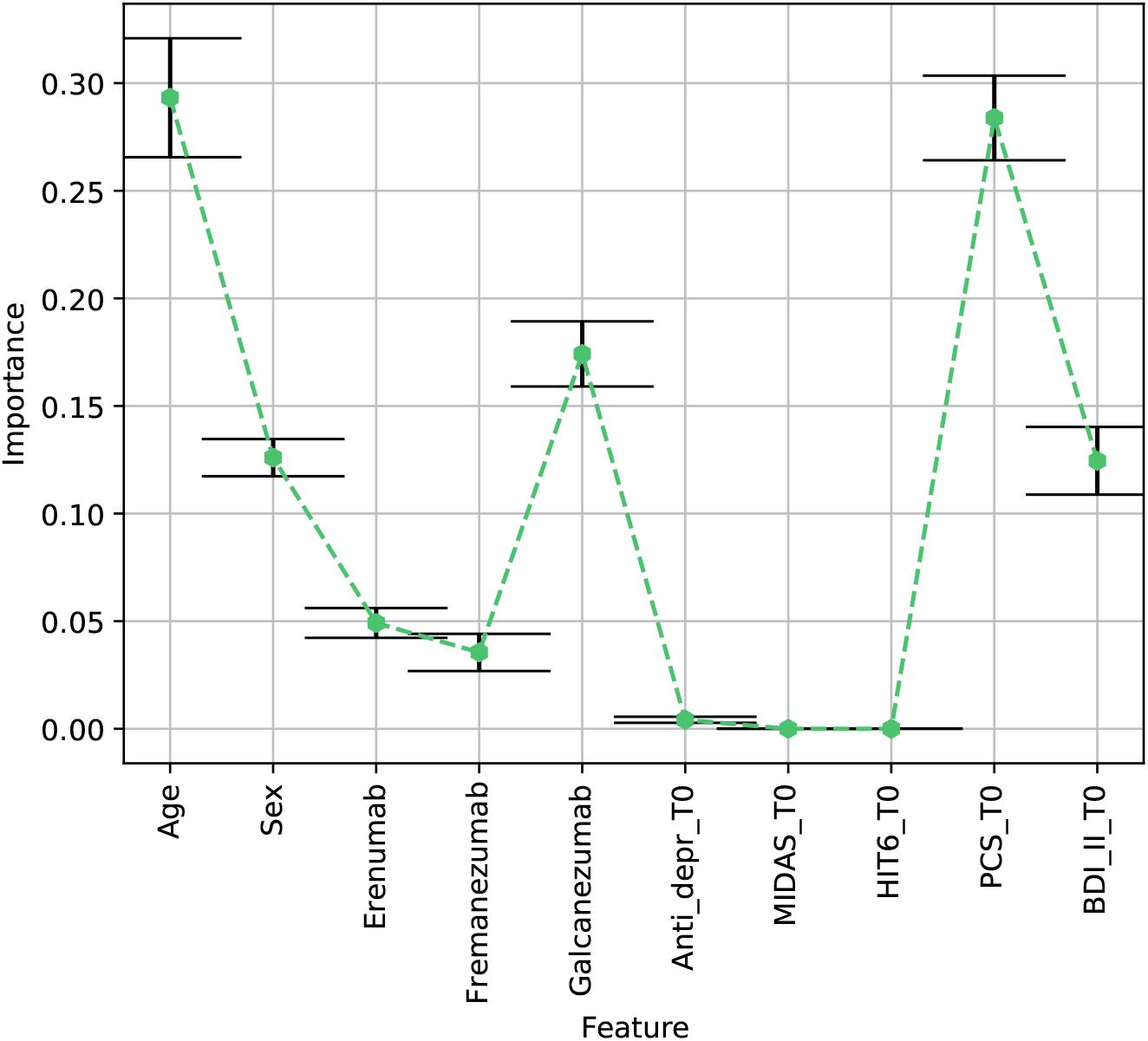
Mean Importance (with 95% CI) per each feature utilized in the logit prediction model. On the x-axis the features involved; note that the tick “MIDAS_*T*_0_” refers to a binary variable derived by selecting MIDAS values greater than 11 at *T*_0_. Likewise, “HIT6_*T*_0_”, “PCS_*T*_0_”, and “BDI_II_*T*_0_” refer to binary variables at *T*_0_ with cutoff, respectively, 50, 30, and 13. On the y-axis the mean importance values are reported; when appreciable, confidence bars are also reported.

### E Sample Size

The estimation of the size was aimed to confirm the primary outcomes of our investigation. We recall that we were interested in exploring through Spearman’s test a possible correlation between PCS and MIDAS as well as PCS and HIT 6 at time points *T*_0_, *T*_1_, and *T*_2_.

The formulas provided by May and Looney (2020) represent the key to determining the required sample size for inference based on Spearman’s test. To apply this method, it is required to specify the punctual values of both the null and alternative hypotheses. Therefore, we assumed to null hypotesis to be *H*_0_ : *ρ*_0_ = 0, and the alternative hypotesis *H*_0_ : *ρ*_1_ = 0.65. That is, we want to investigate no correlation (*H*_0_ : *ρ*_0_ = 0) against a moderate (or even stronger) correlation, i.e., *ρ*_1_ = 0.65. Thus, the sample size *n* is given by

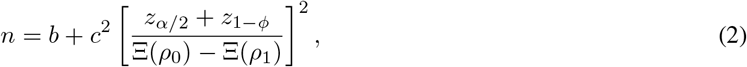

where *b* and *c* are constants taking values 3 and 1, respectively; *α* is the significance level and *β* the test’s power; we used Ξ(·) to denote the hyperbolic arctangent. Note that a regular choice of *α* = 0.10 needs to be adjusted to avoid incurring an increase of Type I errors due to multiple testing. To achieve this, we utilized the Bonferroni correction, so we adopted *α* = 0.0125; see appendix A. We also chose a high-power test to exclude the presence of Type II error, so we opted for *ϕ* = 0.8.

We considered the possibility of dropouts during the phase of data acquisition. Thus, we adjusted the sample size through the following formula

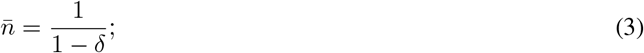

where *δ* denotes the *dropout rate*; we opted for *δ* = 0.15. Applying both (2) and (3) we obtained a population size of 25.

## F Randomization

We implemented a *simple randomization* Altman and Bland (1999) scheme to assign antibody therapy to participants. This method ensured that each participant had an equal probability of receiving the antibody therapy, thereby minimizing selection bias and ensuring a balanced distribution of treatments across the study groups. As outlined in the main manuscript, Erenumab was assigned to 6 patients, Fremanezumab to 7 patients, and Galcanezumab to 12 patients. Despite assigning Galcanezumab to the majority of patients, the number of doses assigned resulted in being equally likely according to the chi-squared test.

We aimed to test the null hypothesis that the antibodies were assigned to patients with equal probability. As described, we utilized the chi-squared test with two degrees of freedom, setting a significance level of *α* = 0.05. The test yielded a χ^2^ statistic of 1.625 and a p-value of 0.4437. Given these results, we do not have sufficient evidence to reject the null hypothesis, indicating that the distribution of antibody assignments is consistent with equal probability.

### G Scales

MIDAS measures the headache-related disability within the last 3 months in three areas of life: work/school, household chores, and family/social/leisure activities. The MIDAS score is formed by summing the items and ranges from 0 to 270. The disability can be categorized into four grades: (I) little or no disability (MIDAS score 0–5), (II) mild (6–10), (III) moderate (11–20), and (IV) severe disability (> 21) (Stewart et al., 2001b).

HIT 6 is a six-item self-reported questionnaire used to assess headache-related disability. It assesses headaches’ impact on psychological, cognitive, occupational, and social functioning over the previous four weeks. Scores range from 36 to 78, and scores above 60 indicate that headache seriously impacts functioning (Rendas-Baum and et al., 2014).

PCS was used to measure pain-related catastrophic thoughts. It consists of three subscales: helplessness (Qs 1-5 and 12), magnification (Qs 6, 7, and 13), and rumination (Qs 8-11). Evaluations (0 = not at all to 4 = always) are made using a five-point Likert scale. The PCS is rated from 0 to 52 points, and a higher score corresponds to a higher level of pain-related catastrophic thoughts(Darnall and et al., 2017).

The Beck Depression Inventory-II (BDI II) is a 21-item assessment of depression’s severity. Each item is graded between 0 and 3 on a four-point Likert-like scale. Higher scores indicate more severe depressive symptoms (Schotte et al., 1997b).

## Data Availability

All data produced in the present work are contained in the manuscript

## Acknowledgments

The authors would like to acknowledge the “Azienda Sanitaria SS. Antonio e Biagio e Cesare Arrigo” for their support.

## Declaration of generative AI in scientific writing

The authors declare that no generative AI and AI-assisted technologies have been utilized during the writing process of this manuscript.

